# Turning Challenges into Opportunities: Lessons from Ethiopia’s COVID-19 Response for Strengthening Health Systems and Health Security

**DOI:** 10.1101/2025.03.17.25324158

**Authors:** Muluneh Yigzaw Mossie, Jenny Shannon, Amare Bayeh Desta, Etsub Brhanesilassie, Adugna Dufera, Saira Nawaz

## Abstract

The COVID-19 outbreak, which emerged in late 2019, rapidly evolved into an unprecedented global public health crisis. An assessment of Ethiopia’s responses to the COVID-19 pandemic offers a unique learning experience for building stronger health systems and enhancing resilience against future health threats. This paper covers the major findings of a national study of the Government of Ethiopia’s COVID-19 response. The study followed a concurrent nested mixed methods design, combining quantitative and qualitative data collection at all levels of the health system. Primary data were collected from surveys of individuals involved in the COVID-19 pandemic response, health facility assessments, key informant interviews, and a review of COVID-19 response norms and standards. The study revealed that Ethiopia implemented a wide range of interventions across the WHO response pillars including multi-sectoral coordination and actions, partnerships with communities by engaging public figures, expanding molecular laboratory capacity, strengthening critical care facilities, and enhancing digitalization of health data. However, we detected a decline and a lack of institutionalization of COVID-19 practices, particularly in the areas of surveillance, laboratory testing, and case management. Our findings thus suggest that COVID-19 investments led to a strong response with robust implementation, but in the absence of a strong transition plan and limited capacity to apply the lessons learned, many of the COVID-19 response platforms were not effectively leveraged to strengthen the health system to address future responses.

## Introduction

The COVID-19 outbreak, which emerged in late 2019, rapidly evolved into an unprecedented global public health crisis [1–3][4]. By the end of April 2023, when the World Health Organization (WHO) declared that COVID-19 was no longer a public health emergency of international concern, more than 765 million people worldwide had contracted the virus, and over 6.9 million had died [5].

Despite causing a significant crisis, COVID-19 offered a unique learning experience, exposing weaknesses in international collaboration but also igniting a surge of global health initiatives and scientific breakthroughs, including the unprecedented rapid development of vaccines [6]. Many countries adapted the WHO COVID-19 response pillars to contain the virus’s spread and manage severe cases. These pillars encompassed coordination and planning, risk communication and community engagement, surveillance, points of entry, laboratory testing, infection prevention and control, case management, logistics and supply systems, maintaining essential services, and research and innovation[7]. The pillars align closely with broader health system components, particularly primary health care systems. The integration and institutionalization of these interventions within the broader health system have thus significant implications for health system strengthening and resilience. This, in turn, reinforces health security, as robust health systems are essential for ensuring health security.

The critical role of health system strengthening in achieving health security and resilience is well-documented [8]. Everyday resilience is essential for maintaining continuity in essential services and for effectively preventing and mitigating emergencies [9]. Strong, adaptive health systems are crucial for sustaining quality care, enabling agility and responsiveness during both routine operations and emergencies[8,10]

### COVID-19 in Ethiopia

Ethiopia reported its first laboratory-confirmed COVID-19 case in March 2020, at a time when the country was not fully prepared [11]. There was no in-country laboratory testing capacity, and human resources and supplies were insufficient to meet the surge in demand [12]. Despite these challenges, the Ethiopian government, in collaboration with its partners, implemented a national response in line with WHO response pillars, specifically establishing multi-sectoral and multi-stakeholder coordination mechanisms, mobilizing resources and financing efforts, fostering partnerships and engaging the private sector and communities, training and deploying surge teams, and strengthening contact tracing and isolation systems [13]. Moreover, efforts focused on improving data quality and utilization for decision-making, enhancing surveillance at points of entry, expanding testing facilities, boosting case management capacity, and increasing vaccination coverage were the key interventions [14].

This study, conducted in Ethiopia shortly after the WHO declared that COVID-19 is no longer a public health emergency of international concern, investigated the strategies used to contain the pandemic, the enabling conditions for the implementation of the response, the barriers encountered, the successes, and the lessons learned to strengthen health care systems for health security. The study aimed to systematically assess what worked well, document experiences and lessons, and leverage these insights to strengthen everyday system resilience and national health security.

## Methods

### Study framework

This study employed two widely recognized frameworks to assess the implementation of Ethiopia’s COVID-19 response. First, an adapted version of the Consolidated Framework for Implementation Research (CFIR) [15] was applied to examine the internal and external contextual factors influencing COVID-19 response implementation in Ethiopia. Originally developed in 2009 and refined over time, the CFIR has been widely applied to explore the facilitators and barriers of implementing various interventions. Second, the WHO COVID-19 Monitoring and Evaluation Framework[7] was employed to synthesize and capture the success and key learnings for each response pillar.

### Study Design

The national COVID-19 assessment in Ethiopia used a concurrent nested mixed methods design, combining quantitative and qualitative data to investigate various pillars of the response. Data were collected from surveys and interviews of individuals involved in the COVID-19 pandemic response, health facility assessments, and a review of the COVID-19 response norms and standards.

### Study Sample

The respondent universe included individuals engaged for 12 or more consecutive months in implementing pandemic response activities at national and/or subnational levels including designing, implementing, and monitoring various aspects of the COVID-19 response. Study participants included those who completed a survey about the implementation process, and a subset of survey respondents that participated in key informant interviews.

At national level, sampling was conducted in two stages. In the first stage, organizations involved in the COVID-19 response were purposively selected. The sample included government organizations such as the Ministry of Health, EPHI, the Ethiopian pharmaceutical supply service, and the Ethiopian food and drug authority. In addition, the sample encompassed donors, international non-governmental organizations, and professional associations.

In the second stage, individuals within these organizations were selected based on their involvement in the COVID-19 response. These aspects included policy formulation, coordination, resource mobilization and financing, surveillance and outbreak investigation, points of entry, laboratory testing, case management, community engagement, infection prevention and control, COVID-19 vaccination, maintaining essential services, data and innovation, and supply systems.

### Sampling of regions

At sub-national level, we sampled 5 of the 13 regional states and city administrations for two predominantly agrarian regions, one city administration, and two predominantly pastoralist regions were chosen. Regional-level organizations that coordinated the COVID-19 response were then purposively selected.

### Sampling of *woredas* and health facilities

*Woredas* within the regions were randomly selected from the list of *woredas* in the DHIS2 database, and subsequently, health facilities were chosen within these selected *woredas*. In most *woredas*, only one health facility providing COVID-19 treatment and isolation, was included in the sample. In *woredas* where two or more facilities offered comprehensive COVID-19 services, one was randomly chosen.

### Sampling of study participants at sub-national level

Organizations at sub-national level were first selected, followed by the selection of study participants. As Figure 1 shows, individuals directly involved in the COVID-19 response for at least 12 consecutive months were chosen and invited for interviews. If an organization or health facility had two or more individuals with the same level of experience, one was randomly selected. Similarly, if an organization implemented multiple COVID-19 response pillars, and different individuals were responsible for specific interventions, all of them were invited for interviews.

**Figure 1:**
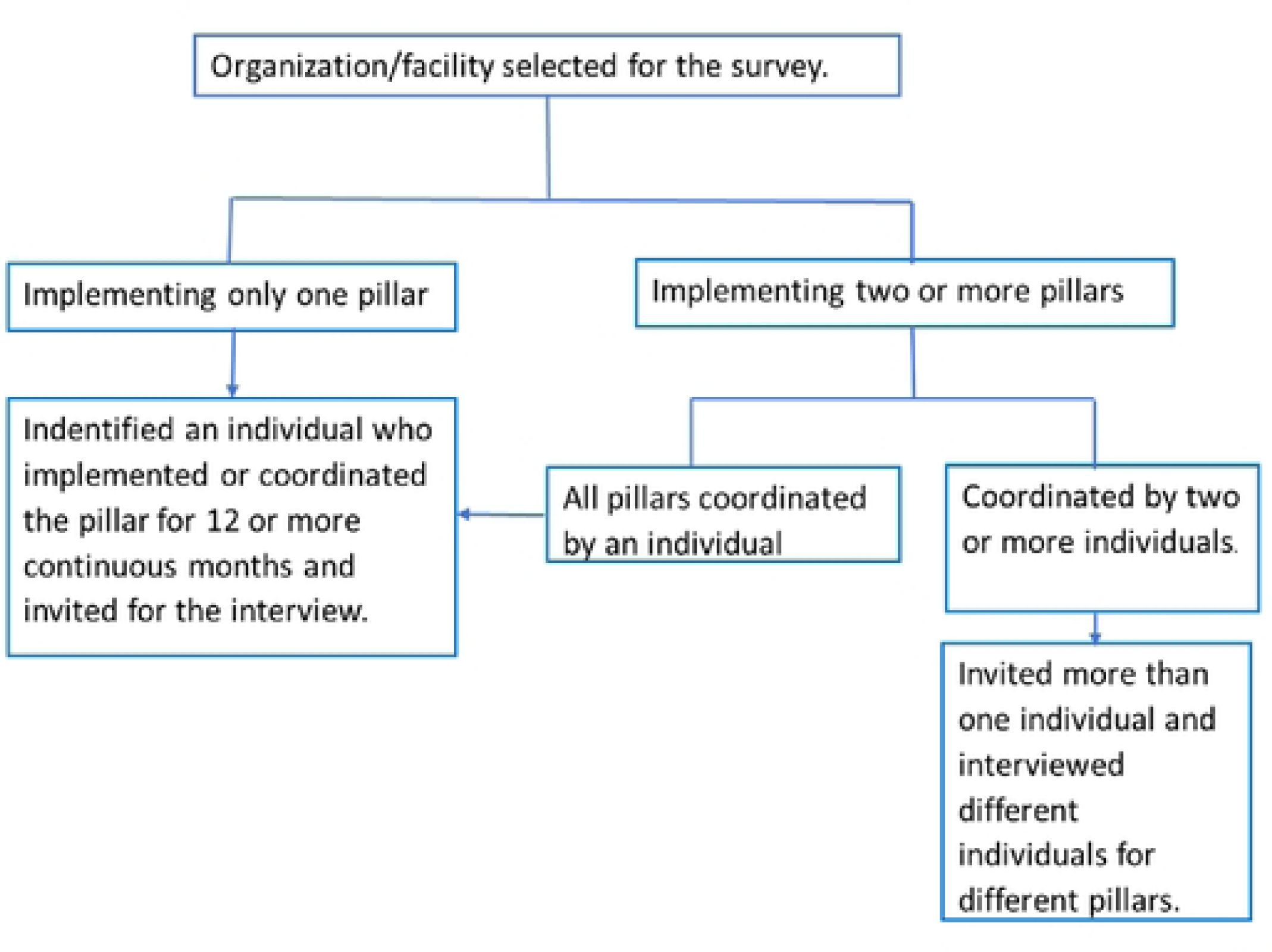
Study participant selection process within organizations.

### Data Collection tools

#### Implementation survey

Using a structured questionnaire, individuals who had experience in policy design, strategy formulation, and implementing COVID-19 response strategies in Ethiopia were surveyed on various aspects of the response, focusing on the enablers and challenges encountered. Insights were gathered from individual, organizational, political, economic, and technological perspectives to capture a comprehensive view of the factors that supported or hindered response efforts. This approach aimed to identify what worked well and what did not, offering a broad understanding of the complexities involved in managing the pandemic within Ethiopia’s unique context.

#### Health Facility Assessment

Respondents from health facilities were asked to reflect on their experiences in developing the capacity of their facilities, including health workforce deployment, training, data recording, reporting and analysis, infection prevention and control within health facilities, critical care capacity, surveillance, and laboratory testing.

#### Key informant interviews

Key informant interviews were conducted with a nested sample of individual survey respondents selected to ensure representativeness across pillars, regions and woredas. Key informants were asked in-depth about pillar-specific implementation strategies, challenges encountered, successes of implementation, and the insights gained during the response.

### Analysis

Data analysis was conducted at three levels: quantitative analysis, qualitative analysis, and synthesis and triangulation. Data from the facility survey was analyzed using descriptive analytics of the level of adoption of guidelines and practices advanced at the health facility level.

For the implementation survey, enablers and barriers to implementation of the various pillars were identified within the health system environment, political environment, economic environment, social environment, and technological environment.

For the qualitative data from KIIs, an initial codebook was developed based on the study questions, the WHO COVID-19 monitoring and evaluation framework, and the interview guide. Upon review of the transcripts from KIIs, the codebook was further refined, and line-by-line coding was performed. The analysis employed both deductive and inductive approaches, identifying common themes based on recurring topics, ideas, and patterns. Atlas.ti, a qualitative data analysis software, was used for the analysis.

Finally, the quantitative and qualitative data, along with information from other documents, were triangulated to inform the development of the results. Data from all sources were synthesized to better understand the variability in program rollout across regions and woredas and to explore how contextual factors facilitated or hindered implementation.

We assessed the robustness of the evidence supporting key findings based on three dimensions: the level of data (whether multiple or a single data source was used), the type of data (whether it was perception-based or fact-based), and the quality of the data.

### Ethical considerations

The Ethiopian Public Health Institute Institutional Review Board approved the study (Protocol Number: EPHI-IRB-512-2023). Recruitment of study participants took place from August 7, 2023, to December 15, 2023, in accordance with ethical guidelines. As per IRB approval, verbal consent was obtained from all participants after they received and read the consent form, which detailed the study’s purpose, potential benefits, risks, and possible harms. Participants provided verbal agreement to proceed with the interview, and the interviewer documented consent by marking a designated section on the form. All completed consent forms were securely retained to ensure proper ethical documentation and compliance. Additionally, all data were deidentified and stored on a secured, password-protected device to maintain participant confidentiality and data security.

## Results

Ethiopia implemented a wide range of interventions across the WHO response pillars. Measures such as contact tracing, isolation, and quarantine at points of entry were implemented beginning from the early phases of the pandemic. In addition, Ethiopia leveraged digital tools for data collection and analysis, enabling surveillance and monitoring of the pandemic. Laboratory testing and case management were scaled up to identify, isolate and treat cases, while efforts were made to ensure a steady supply of essential medical tools. In the later phases of the pandemic, the government-initiated vaccination programs to curb the virus’s impact further. The government established a range of directives and regulatory frameworks to facilitate the implementation of these interventions, particularly during the initial phase of the pandemic, including the declaration of a state of emergency. A key informant from EPHI described the government’s role in establishing legal frameworks as a key facilitating factor for the entire response:

> “From the outset, laws, rules, and regulations were issued frequently. Policies were introduced requiring individuals to maintain physical distance, wash their hands, and ensure that hotels, schools, and other crowded service facilities operated in compliance with these regulations. Adjustments were made as necessary based on evolving circumstances. If the government had been unable to establish such regulations, implementing the [COVID-19 response] interventions would have been difficult.”

> -A respondent from EPHI

### Multi-sectoral and multi-stakeholder collaboration and action

Multi-sectoral and multi-stakeholder coordination mechanism was established at the first phase of the COVID-19 pandemic with higher political engagement. The taskforce, which was established through the multi-sectoral and multi-stakeholder coordination mechanism, rapidly devised new legal and regulatory provisions, outlining the dos and don’ts of non-pharmacological interventions nationwide. A key informant from the Ministry of Health noted:

> “Many stakeholders were involved in the COVID-19 coordination. These included the EPHI team, the ministry of health team, and other stakeholders. When the response plan was prepared, we involved not only the public health institute, but also various other stakeholders.”

> –A respondent from the Ministry of Health

There were challenges in the coordination mechanisms specifically at the early phase of the pandemic. Conflict compromised the implementation of the COVID-19 national and sub-national responses, as the public failed to honor directives and government officials were unable to proactively involved in the taskforce as they prioritized other public demands. Furthermore, following the war in northern Ethiopia, the government’s focus shifted to security, resulting in little engagement in the COVID-19 response. In the Amhara region, a respondent specifically highlighted the significant impact of the war on the COVID-19 response efforts within the region.

> “…There was a conflict in northern Ethiopia…and during that time, high-level officials were seen chatting with military personnel in crowded conditions. People observed them without face masks in a meeting hall filled with many individuals. Consequently, the public misperceived that COVID-19 was not present in the country.”

> –A respondent from a regional public health institute

Despite challenges and setbacks in the coordination mechanism, the Ministry of Health and its partners developed strategies to contain the pandemic, even in conflict-affected settings, with a particular focus on revitalizing vaccination efforts. Coordination and partnerships among stakeholders enabled the vaccination of notable number of people, even in these challenging areas. A key informant at sub-national level said:

> “…to reach hard to reach areas, we have been using a mobile approach, where a mobile team is assigned to reach each cluster. We tried to serve IDP sites and refugee camps using this method. The funds required to establish the mobile teams were found by coordinating different partners. There are also, hard to reach places …, places not reachable due to security reasons, where we followed a hit and run approach to reach them.”

> - A respondent from a zonal health department

### Training and deployment of Rapid Response Teams

Soon after the World health Organization declared that COVID-19 is a public health emergency of international concern, the Ethiopian Ministry of Health established and revitalized RRTs. These teams were composed of epidemiologists, clinicians and WASH experts, risk communication and community engagement experts, laboratory personnel, and logisticians. The roles of RRTs were wide-ranging, covering nearly all aspects of response except for vaccination, which was managed by the Expanded Program on Immunization (EPI) unit at the Ministry of Health, independently of RRTs. Stationed at the national and regional Emergency Operating Centers (EOCs), RRTs were involved in preparing suspected cases for testing, facilitating their isolation when necessary, and ensuring their admission to case management centers as appropriate. A key informant described the roles of RRTs as:

> “…The primary component [of the COVID-19 response activities] is what we refer to as the rapid response team, which was deployed in all kebeles, the lowest level of administration in the country. So, the rapid response team was not solely focused on response activities but also conducted searches for suspected cases.”

> -A respondent from the Ministry of Health

### Contact tracing, quarantine and isolation

At the first phase of the pandemic, all suspects and travelers were quarantined until confirmed negative. Those found positive for the virus were also isolated regardless of the severity, although later home-based management became common practice rather than isolation in designated centers. Quarantining and isolation of enormous number of people – all contacts, travelers, and positive cases-created a challenge in a health system which was already resource constrained. Inadequate supplies at quarantine and isolation centers hampered the capacity to safely isolate and manage all these suspected and confirmed cases. In addition, contact tracing was challenging due to deficits in communication skills among the RRTs as their training in interpersonal communications was limited due to an emergency deployment, hindering their ability to effectively engage with contacts. Adding to this, inadequate resources at isolation centers exacerbated hesitancy among suspected cases to cooperate and adhere to isolation and quarantine measures. A key informant remarked

> “Managing a large number of people … proved to be quite demanding. Despite the efforts of various partners to supply food and water, meeting the needs of all individuals was challenging. Moreover, the warm weather conditions contributed to some individuals escaping from the quarantine center and rejoining society.”

> -A respondent from a zonal health department

To overcome resource constraints, the federal and regional governments undertook resource mobilization efforts, largely by engaging local communities, the private sector, and donors. Higher learning institutions also contributed by providing student facilities for the quarantine of returnees. A key informant from one of the regions noted:

> “The private sector contributed a lot in funding the [COVID-19] response, mostly in kind. We conducted meetings with industry managers and wealthy people living in the town. …After that, local businesses supported us with a lot of money. … The textile industry produced many masks. Private health facilities collected money and brought us thousands of masks.”

> -A respondent from a regional health bureau

### Availability and quality of data for surveillance

When the COVID-19 pandemic struck, there was an absence of reliable databases for surveillance and outbreak investigation in Ethiopia. Surveillance personnel resorted to manual recording of cases, followed by the transfer of data to higher levels using available open applications. This approach resulted in errors in both recording and reporting, contributing to incomplete data. In addition, reporting materials ran out of stock, forcing surveillance personnel to search for copies while also dealing with time-consuming and complex paper forms. However, as the pandemic progressed into mid-2020, a notable milestone was the development of an integrated application suite. This application system incorporated surveillance, laboratory testing, point of entry, case management, and logistics information systems. Leveraging the DHIS2 platform, the application integrated all pillars and established interfaces to facilitate communication and data driven decision making among all responders involved in the COVID-19 response efforts. Two key informants explained the initial challenges and the role digitalization played in the COVID-19 response:

> “There was a system called DHIS2 for [real time recording of] COVID-19 that allowed us to register directly on the system. And we have access to the data from there…for this reason, the reports were immediate and no delay. The application allows professionals at zone and woreda levels to analyze the data and generate information for action.”

> -A respondent from a regional health bureau

> Initially, the surveillance and data capture system relied on Excel, leading to issues with missing, inconsistent, and untimely data. However, the Ministry of Health leadership later prioritized digitalization, resulting in a fully digital system with integrated data interfaces, streamlining data from various sources.

> -A respondent from EPHI

### Surveillance at point of entry

Ethiopia established a network of 27 points of entry to monitor and control the influx of travelers during the COVID-19 pandemic. Respondents believe that despite being absent at an early phase of the pandemic, the deployment of surge teams at points of entry boosted response capabilities, allowing for rapid actions in the event of a surge in arrivals or suspected cases. A key informant at subnational level described:

> “We had dispatched a surge team to the area [point of entry] to conduct screening for infectious diseases, including COVID-19. It’s important to note that the displaced individuals arriving from the conflict in Sudan included not only Ethiopians but also individuals of other nationalities.”

> –A respondent from a regional health bureau

Although Ethiopia set up a network of point-of-entry surveillance systems, porous borders and limited cross-border collaboration remained significant challenges. The absence of cross-border surveillance mechanisms further impeded effective outbreak investigations. In areas lacking formal checkpoints, people crossed the borders unchecked, mingling with nearby communities and breaching prevention guidelines.

### COVID-19 laboratory testing

When the COVID-19 pandemic occurred, Ethiopia’s laboratory testing capacity was limited. In February 2020, only one laboratory in the country was capable of performing COVID-19 RT-PCR testing. Moreover, the absence of automation and the prolonged turnaround times for test results presented challenges during the early stages of the pandemic. Two respondents explained the state of laboratory services when the pandemic occurred in Ethiopia:

> “Initially, we only had one molecular laboratory at a national level, located at EPHI, the Influenza lab. We didn’t even have a molecular laboratory professional.”

> -A respondent from the Ministry of Health

> “Initially, due to outdated methods and a limited number of professionals, it took over 24 hours to process samples. Moreover, with all samples from different regions being handled by a small team at the national lab, accessibility to laboratory tests was impacted.”

> -A respondent from EPHI

Despite these initial setbacks, as the pandemic evolved, Ethiopia was able to expand testing centers. The country conducted an inventory of available PCR machines, including those in research centers across sectors like agriculture and higher education. Collaborative platforms were established with these organizations, enabling the repurposing of existing machines and enhancing testing capacity. Moreover, private laboratories in Addis Ababa played a role in expanding testing facilities by offering COVID-19 RT-PCR testing, particularly for travelers. Consequently, the number of laboratories equipped with RT-PCR testing capacity increased from just one in February 2020 to 72 by June 2020 (Figure 2).

**Figure 2:**
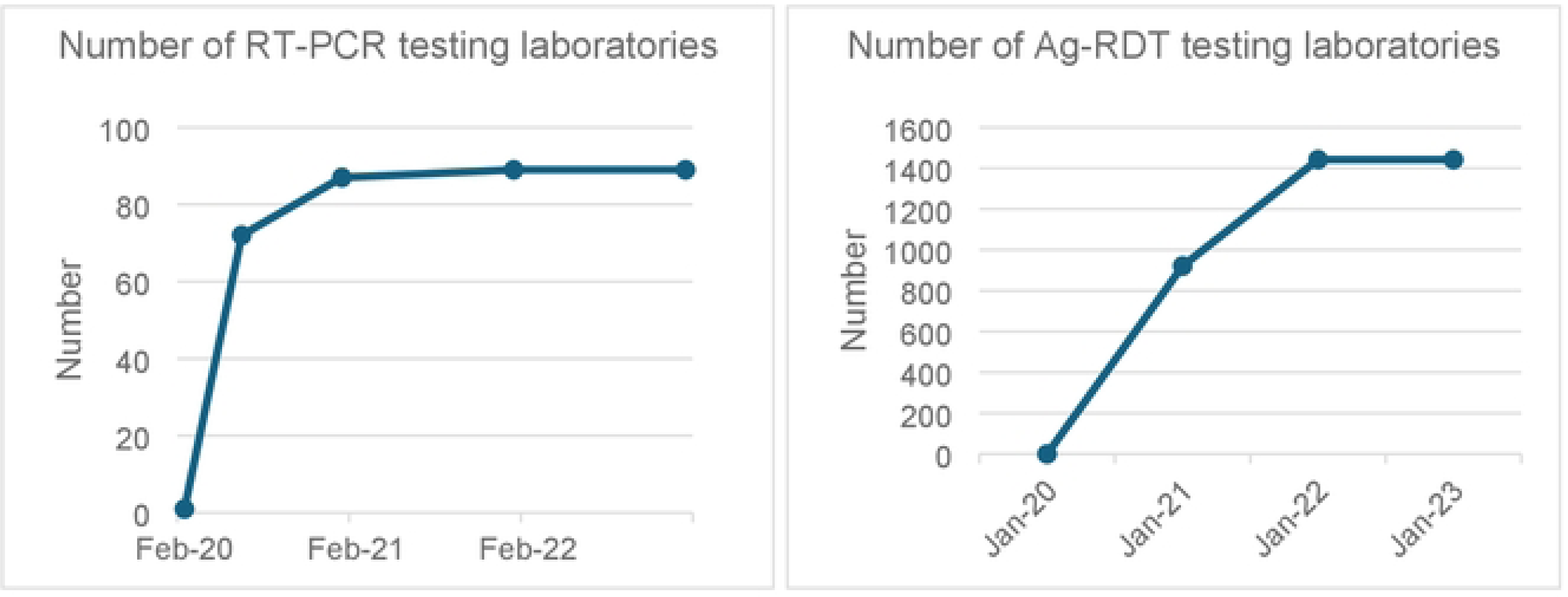
Trends in COVID-19 laboratory testing capacity during the pandemic period.

In addition, Ethiopia was able to conduct in-country viral genome sequencing, enabling the country to monitor evolution of the virus. A key informant noted the following:

> “…Initially, we didn’t have a capacity for viral genome sequencing. So, tracking the variants was a challenge. I think it’s still a challenge, but at least we now have in-country capacity. This capability is available at the national level, primarily at EPHI and a few other laboratories. So, identifying which variant is responsible for a surge in a specific area can now be determined locally. That is a significant development.”

> -A respondent from the Ministry of Health

In the middle of the pandemic, Ethiopia approved and deployed antigen-based rapid diagnostic tests (Ag-RDTs) in areas where RT-PCR testing was not available. By 2022, 1,442 health facilities were conducting Ag-RDT testing for COVID-19. Over time, the automation of laboratory processes further enhanced daily testing capacity. The advent of rapid diagnostic tests and expansion of RT-PCR testing centers improved access to laboratory testing. In 2021, close to 78% of case management and isolation centers had SOPs to conduct onsite COVID-19 testing, while it steadily declined there after (Figure 3).

**Figure 3:**
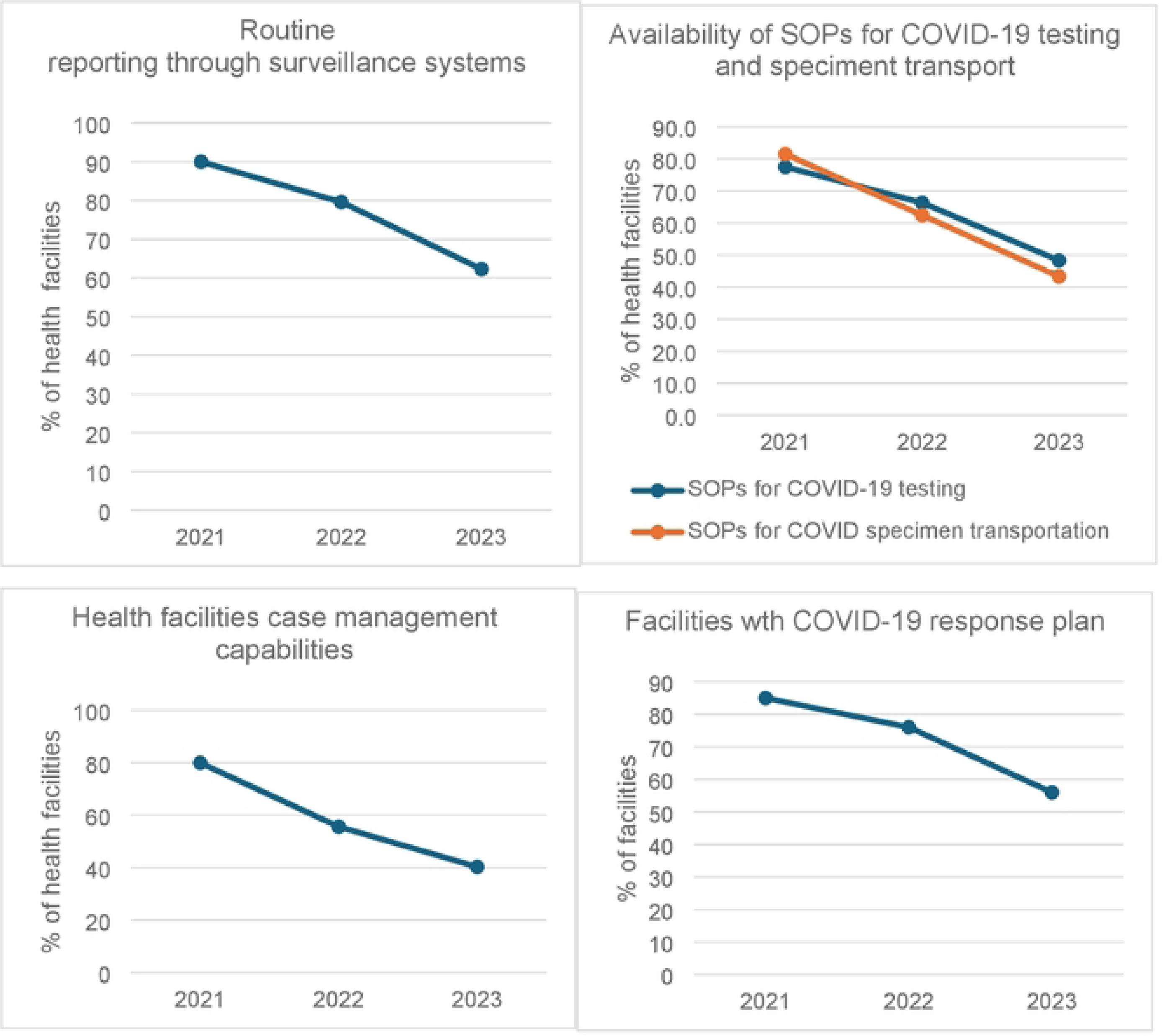
Trends in implementing key intervention in health facilities from 2021 to 2023.

Although the sharp increase of testing center for COVID-19, the infrastructure of laboratories remained unchanged, resulting in insufficient space for testing activities. This led to overcrowding and congestion within the laboratories, increasing the risk of sample mixing and contamination. A key informant described the limited spaces available for laboratory testing and the challenges of quality assurance of laboratory testing:

> “…the infrastructure in the laboratories did not meet the standard for a molecular lab. … Some regions simply provided one room and expected us to work there. However, this was difficult because contamination is a significant challenge in molecular samples and could even result in false positive results”

> -A respondent from EPHI

In addition, ensuring access for all populations to laboratory services remained challenging due to Ethiopia’s vast geography and the growing number of COVID-19 cases. To address access issues in remote areas without testing centers, samples were collected and transported using existing infrastructure. Ethiopia’s COVID-19 testing strategy utilized the country’s established systems and experiences for sample collection, transport, and testing, which were already in place for diseases such as tuberculosis, HIV, and sentinel surveillance of Severe Acute Respiratory Illness and Influenza-like illnesses.

### Case management

When the COVID-19 pandemic hit, Ethiopia had little capacity to manage severe cases of COVID-19 as all key informants noted. The country was grappling with a lack of trained health providers, ventilators and other essential equipment and supplies for critical care, although case management centers were established early in the pandemic. To address this, an online training platform was established, and several health providers trained remotely.

> “We organized several training sessions on COVID-19 for healthcare workers at the early phase of the pandemic, which served as a crucial starting point since it was a new disease and there was limited knowledge on how to respond. To address this, we established an online training platform that enabled us to train many health workers in a short period. We also created a central information repository, compiling the latest guidance and publications about COVID-19. This repository was made accessible through a dedicated platform.”

> A respondent from a professional association

By the end of 2021, the number of case management centers established nationwide reached 155. With the expansion of case management centers, the number of critical care devices also grew. Throughout the pandemic, Ethiopia secured over 764 oxygen concentrators and 320 mechanical ventilators (Figure 4).

**Figure 4:**
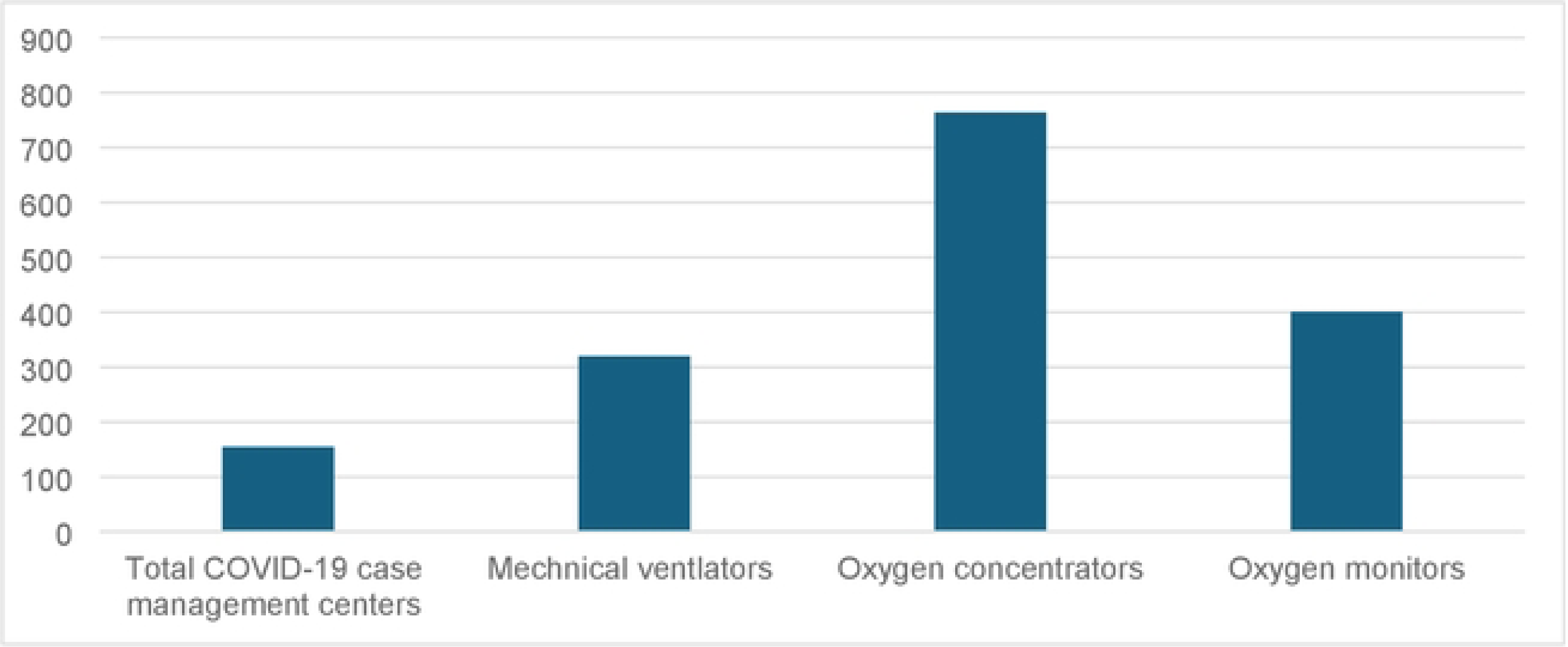
Total number of COVID-19 case management centers, mechanical ventilators, oxygen concentrators and oxygen monitors secured during the pandemic.

A key informant from the Ministry of Health described the improvements in critical care facilities as follows:

> “…the case management has changed a lot of things, for example, our oxygen system has been changed, beds were not connected to oxygen in any of the hospitals before the COVD pandemic. In addition to the oxygen plant, oxygen cylinders have been procured in large quantities. Mechanical ventilators were few before COVID, but now we have many.”

> -A respondent from the Ministry of Health

As case management centers expanded, the inadequate availability and unsuitability of personal protective equipment (PPE) jeopardized the safety and well-being of frontline health workers, increasing their risk of contracting the virus while treating patients. This shortage also heightened the risk of COVID-19 transmission within health facility premises, further disrupting essential health services in both case management centers and other facilities. In the later stages of the pandemic, improvements in PPE availability and usage, along with increased awareness among health workers and the public, led to a reduction in disruptions to essential services (Figure 5).

**Figure 5:**
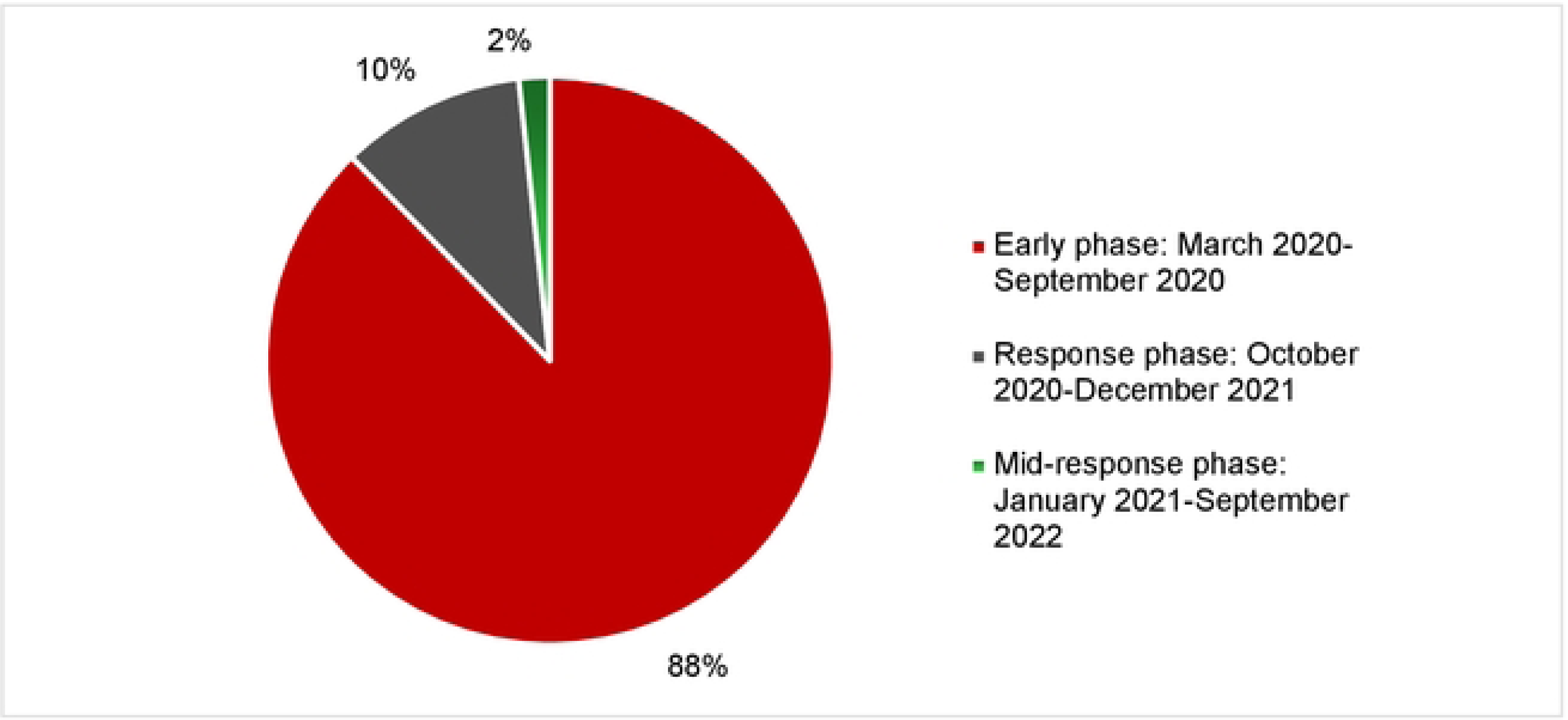
The phase of the COVID-19 pandemic when the most disruption to the health system was observed.

### Logistics and supply systems

A subcommittee under the national COVID-19 task force was established to coordinate logistics and supply availability, focusing initially on items such as PPE, mechanical ventilators, laboratory devices, and, later, vaccines. This committee, composed of members from the Ministry of Health and other sectors, played a role in ensuring efficient resource distribution.

Initially, as the pandemic created global demand, widespread shortages of supplies emerged in international markets. The persistent scarcity and rising costs of PPE imposed financial burdens on both the government and the public in Ethiopia. In response, the logistics and supply chain task force explored local production options, collaborating with local companies with manufacturing experience. Through these public-private partnership efforts, local production capabilities were rapidly mobilized, leading to the production of essential supplies within a month and at affordable prices. A key informant explained how development of local production capacities improved cost and availability of face masks:

> “During the early stages of the pandemic, the price of a single face mask skyrocketed from its usual 5 birrs to as high as 350 birrs. However, within a month or even less, we successfully ramped up production and imports of masks, consequently stabilizing the market. This achievement directly resulted from the strategies we put in place and stands as one of the measures, among others, that contributed to our improved control over the pandemic.”

> -A respondent from the Ministry of Health

Coordination and collaboration among sectors played a role in expediting the last mile delivery of logistics. By working closely with the Ministry of Finance and Customs, health authorities at the Ministry of Health and regional health bureaus were able to enhance the distribution and timely access to essential items required for the response. New directives were also established to facilitate the distribution of essential supplies to final destinations within a short period. A key informant described the role of multi-sectoral coordination in the swift transportation and delivery of essential supplies:

> “The coordination across sectors made the logistics to go out quickly in a short period of time and the items were not warehoused… In the past, …commodities imported as emergency used to stay at customs for 15 days and even months, but during the time of COVID-19 pandemic, the commodities arrived [to final destinations] immediately. The Ministry of Finance, Customs, EFDA working together, especially the laws issued by the EFDA, made the COVID-19 response commodities reach the final destination within a short period.”

> –A respondent from EPSA

### Vaccination

The commencement of the COVID-19 vaccination initiative in Ethiopia occurred exactly one year after the country’s first reported COVID-19 case. COVID-19 vaccination started within the established EPI structures and coordination platforms, leveraging their existing functions. These included utilizing pre-existing advisory groups and committees like the Interagency Coordination Committee (ICC) and the National Immunization Technical Advisory Group (NITAG).

Ethiopia aimed to vaccinate up to 20% of the target population in the first phase of its vaccination initiative [16]. However, the country only managed to vaccinate 3% of the target within six months. While limited vaccine stock was a challenge, an ineffective vaccination strategy was also another barrier at the first stage of the vaccination drive. When the initial approach failed to reach target populations, Ethiopia adopted a hybrid delivery strategy with campaign style approach playing a central role, allowing the program to access underserved groups and substantially increasing the number of people receiving vaccine doses (Figure 6).

**Figure 6:**
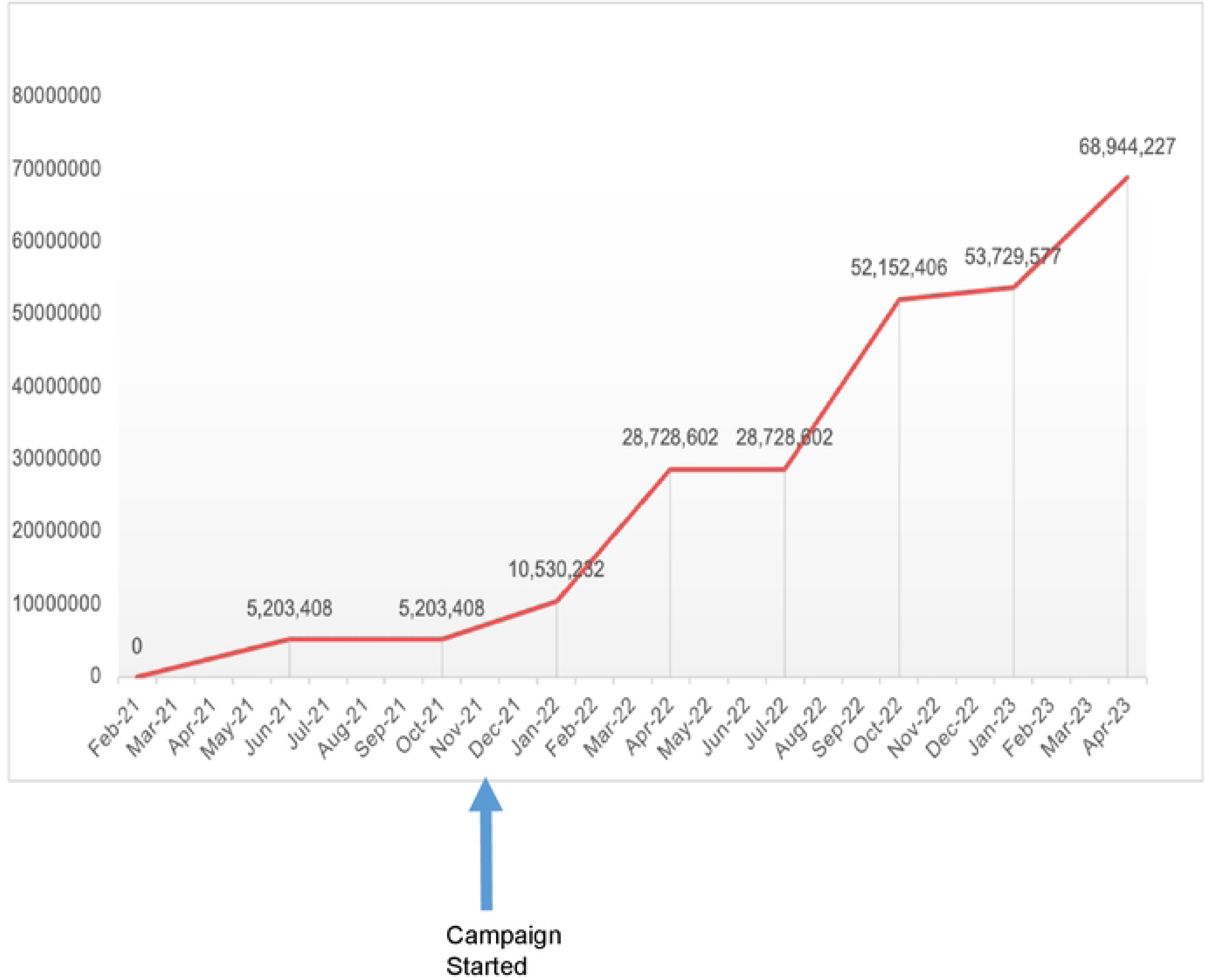
Trends in Covid-19 Vaccine Doses Administered in Ethiopia.

Moreover, Ethiopia enhanced its cold chain infrastructure, particularly the ultracold chain capacity. With support from development partners, ultracold chain facilities were established in all regions of the country, significantly increasing their availability.

### Service integration

The Ministry of Health developed guidelines and began integrating COVID-19 response activities and strategies into the broader health system since 2022. According to these guidelines, all health facilities were expected to incorporate COVID-19 measures into routine services, as the disease had transitioned to an endemic phase. Laboratory testing was to be integrated with existing services, and COVID-19 surveillance aligned with established disease surveillance protocols. However, our analysis revealed a steady decline in COVID-19-related activities within health facilities, even prior to 2023, when COVID-19 was still considered a public health emergency of international concern. As shown in Figure 3, the percentage of facilities equipped with functional lifesaving devices to manage COVID-19 cases dropped from 79.9% in 2021 to 40.3% in 2023. Similarly, the proportion of facilities with standard operating procedures (SOPs) for laboratory testing and specimen transport for COVID-19 declined from 81.5% in 2021 to 43.3% in 2023.

## Discussion

Ethiopia implemented a comprehensive set of interventions to contain the COVID-19 pandemic, despite facing numerous challenges. Key measures included multi-sectoral collaboration and action, point-of-entry screening and quarantine, active case finding and isolation, community engagement, expansion of testing and case management facilities, and later, vaccination efforts. With strong political commitment from the outset, Ethiopia swiftly engaged various sectors and mobilized resources and personnel. Early in the outbreak, the government established 27 points of entry equipped with staff and facilities to screen and quarantine travelers, aiming to prevent the importation and spread of the virus.

At the onset of the pandemic, Ethiopia had limited capacity for laboratory testing and managing severe COVID-19 cases [12]. When the World Health Organization declared COVID-19 a public health emergency of international concern on 30 January 2020, Ethiopia lacked laboratories capable of conducting SARS-Cov-2 RT-PCR tests and had insufficient life-saving equipment to manage severe cases. This was a common challenge among many low- and middle-income countries (LMICs), where health systems were generally weak [17]. In these settings, laboratory testing capacity was low, and critical care facilities were limited in the ability to provide adequate care for severe COVID-19 cases [18].

Despite Ethiopia’s limited laboratory testing and case management capacity, the early implementation of non-pharmaceutical interventions effectively delayed the peak of COVID-19 transmission. This delay provided critical time to expand testing capacity and prepare for severe cases, all without imposing strict lockdowns. Although the first COVID-19 case was reported in mid-March 2020, the peak was delayed by several months, occurring between late August and early September 2020 [13]. Similar experiences were reported in other countries. Robust point-of-entry measures, along with vigilant surveillance, social distancing, handwashing and case isolation [19], effectively delayed pandemic spread, buying time to scale up case management and laboratory services. The role of these interventions in infectious disease prevention and outbreak containment is well-established [20,21] and are foundational to primary health care systems today. Long before antibiotics discovered, the germ theory transformed disease prevention, highlighting the importance of hygiene and quarantine measures—key strategies in managing COVID-19.

Across all pillars, Ethiopia’s COVID-19 response leveraged public-private partnerships and collaboration with various sectors, civil society, donors, and international partners for resource mobilization, planning, and funding. More importantly, the collaboration between the agricultural sector, higher learning institutions, and the health sector went beyond traditional boundaries, enabling shared resources and access to critical services. Notably, the health sector’s collaboration with universities and agricultural research institutions facilitated the rapid scaling up of laboratory testing, as PCR machines from these institutions were repurposed for COVID-19 testing. Furthermore, universities provided student facilities for quarantine and isolation, highlighting the significant role higher learning institutions played during the pandemic. The importance of multi-sectoral and multi-stakeholder collaboration in health is a well-documented fact and forms a core pillar of primary health care systems [22]. Further, public-private partnerships are increasingly recognized for their role in health system development, as shared resources and collaborative efforts help secure essential supplies and foster innovation [23]. The private sector played a pivotal role in Ethiopia’s COVID-19 response, particularly by expanding access to laboratory testing, availing PPE, and other essential facilities, showcasing the important role the private sector can play in public health interventions.

In Ethiopia’s COVID-19 response, public figures played a role in strengthening community trust and promoting adherence to non-pharmaceutical interventions. The involvement of religious leaders and other prominent figures helped build community confidence during a time when misinformation and disinformation were widespread, encouraged broader public engagement, and supported compliance with response guidelines and protocols. Building trust and fostering effective public relations have long been recognized as essential for managing health emergencies and other public health interventions, particularly in primary health care systems [24–26]. Reflecting this, global primary health care guidelines emphasize community engagement, as highlighted in a recent WHO and UNICEF publication on the PHC operational framework [22]. Moreover, evidence shows those countries effectively controlled COVID-19 without strict lockdowns partly relied on strong community trust, underscoring its critical role in disease prevention and outbreak response [19].

A key pillar of the COVID-19 response was the provision of essential supplies, including PPE and, later, vaccines. During the pandemic, scarcity of these items was a challenge. Shortages of these essential items were partly due to global supply chain disruptions and vaccine nationalism [27,28], and partly due to inefficient supply systems which was a characteristic of weak health systems [29]. Global supply disruptions were partly overcome by expanding local production capacity in Ethiopia.

The provision of essential medical supplies is recognized as a foundational pillar in both the WHO’s health system building blocks [30] and the primary health care framework [22]. The availability of such critical supplies is essential for the everyday resilience of health systems as well as in mitigating outbreaks. However, LMICs often face persistent gaps in these supplies. While insufficient financing is a contributing factor, inefficiencies in supply chains—particularly in last-mile delivery—pose significant challenges [29]. The global supply chain issues highlighted by COVID-19 emphasize the need to develop local capacity for producing essential medical supplies. By building local production capacity through public-private partnerships, the availability of essential supplies and self-sufficiency can be improved, contributing to stronger primary health care systems.

Manual data recording initially affected data quality, particularly for COVID-19 surveillance and data-driven decision-making in Ethiopia. However, collaborative efforts improved digitalization across the response, streamlining data for surveillance, laboratory testing, and case management. This rapid digital transformation highlighted the potential of digital health solutions in resource-limited settings and has implications for strengthening health system resilience. Together with previous initiatives, such as the Information Revolution Roadmap [31,32], this experience offers a valuable learning opportunity for the Ethiopian government to advance its health information systems.

### Implication of the COVID-19 response in health system strengthening in Ethiopia

Ethiopia has long experience in primary health care, centered on a community health program led by health extension workers. However, recent studies highlight challenges stemming from fragmentation within the program [33]. Launched in 2003, Ethiopia’s community health program was initially designed to empower communities and families in managing their own health, serving as an entry point into the health system [34]. Over time, this focus on community empowerment has diminished as shifting priorities diverted attention from this core objective, leading to fragmented efforts [33,35]. Ethiopia’s National Health Sector Development Program (HSDP), implemented from the late 1990s to 2015, recognized the community health program led by health extension workers as a flagship component of the health system [33]. However, it did not place significant emphasis on involving public figures as part of its community engagement and trust-building strategy. Instead, community engagement was facilitated through the one-to-five network and the Health Development Army[36] approaches that were not explicitly utilized during the COVID-19 response.

The role of community engagement and multi-sectoral action in Ethiopia’s initial COVID-19 response underscores the value of engaging communities in primary health care and health security. While the Ministry of Health has recently taken steps to optimize the community health program [37], valuable lessons from the COVID-19 response remain. Engaging communities through public figures and revitalizing multi-sectoral action enhances the implementation of health programs [38,39], build public trust, and combined with other measures, support the realization of resilient primary health care systems.

The COVID-19 response offers other valuable opportunities for the primary health care system in Ethiopia to learn and grow. One key takeaway is the training provided to health workers across various aspects of the response, primarily through an online platform. This platform has established a legacy, serving as a resource for continuous professional development in Ethiopia’s health system. Furthermore, the over 3,000 RRTs deployed during the pandemic gained extensive experience in managing various outbreak response activities. If properly documented, these RRTs could be rapidly mobilized in the event of future emergencies, representing a significant asset for the development of the health workforce.

The other positive legacy of the COVID-19 response in Ethiopia includes significant developments in laboratory testing, critical care, and digitalization. The expansion of molecular laboratories and the establishment of viral genome sequencing capabilities enhance the country’s ability to detect and monitor diseases. In addition, the health system secured critical medical devices and deployed ultracold chain facilities nationwide. Combined with experiences in digitalization, these developments, if integrated into routine healthcare practices, could transform Ethiopia’s health system and ensure resilience.

### The need to have an integrated health system framework

Over the past one and half decades, Ethiopia’s health system strengthening efforts have focused on WHO’s six building blocks, while health security efforts have centered on IHR core capacities, often overlooking the role of health system resilience in health security. An additional emphasis on primary health care—although seen as integral to health system development—has further fragmented decision-making, policy, and resource allocation. Some policymakers emphasize health security, downplaying system strengthening, while others champion primary health care as central to health system development[40]. When considering how to integrate COVID-19 response lessons into the broader health system, a key question arises: should these lessons be incorporated into health system strengthening via the six WHO building blocks, the IHR core capacities, primary health care pillars, or all these areas? To harness COVID-19 response lessons and build resilience, Ethiopia needs an integrated, context-informed health system framework that reduces siloed approaches, as the WHO building blocks may not fully address health security needs.

Our analysis showed that while Ethiopia attempted to integrate COVID-19 response experiences into its broader primary health care system for routine practice, the implementation of some of the recommended practices declined even before the pandemic ended. This decline highlights the need for leadership that prioritizes service integration into routine practices, as well as a unified framework to guide implementation and ensure resilience. Without such a framework, these lessons and experiences may not be fully sustained, as system level integration has not materialized, and decision making is fragmented. With fragmented decision-making, oversight and monitoring of these valuable lessons within the broader health system will be limited, hindering efforts to ensure everyday resilience.

### Limitations

There were a few limitations to this study. First, the health facilities selected for the sample were deliberately chosen from those designated as case management and isolation centers, with the purpose of obtaining comprehensive information about the response efforts. Consequently, the findings from these facilities may not fully represent all health facilities across the country. Second, the retrospective nature of the information sought may have introduced recall bias, which could affect the accuracy of the study’s findings. Despite these limitations, the study offers valuable insights into COVID-19 response efforts, with data triangulated from multiple sources to enhance data quality and validity.

### Conclusion

Ethiopia gained extensive experience in community engagement and building public confidence throughout the COVID-19 response. In addition, the Ethiopian health system acquired valuable experiences and tools during the COVID-19 response, particularly in areas such as health workforce development, critical care, digital data solutions, cold chain systems, point of entry, and laboratory testing. If these capacities are integrated into the broader health system—especially within primary health care—and sustained, they have the potential to significantly strengthen Ethiopia’s health system, enhance its resilience in daily operations, and provide a robust foundation for preventing and mitigating future risks. However, we detected a decline and a lack of institutionalization of COVID-19 practices, particularly in the areas of surveillance, laboratory testing, and case management. Our findings thus suggest that COVID-19 investments led to a strong response with robust implementation, but in the absence of a strong transition plan and limited capacity to apply the lessons learned led to many COVID-19 response platforms not being effectively leveraged to strengthen the health system for future emergencies.

## Data Availability

The data supporting the findings of this study are available upon request from the corresponding author.

